# Stimulation of vascular organoids with SARS-CoV-2 antigens increases endothelial permeability and regulates vasculopathy

**DOI:** 10.1101/2021.04.25.21255890

**Authors:** Abdullah O. Khan, Jasmeet S. Reyat, Joshua H. Bourne, Martina Colicchia, Maddy L. Newby, Joel D. Allen, Max Crispin, Esther Youd, Paul G. Murray, Graham Taylor, Zania Stamataki, Alex G. Richter, Adam F. Cunningham, Matthew Pugh, Julie Rayes

**Affiliations:** Institute of Cardiovascular Sciences, College of Medical and Dental Sciences, University of Birmingham, Vincent Drive, B15 2TT, Birmingham, U.K.; School of Biological Sciences, University of Southampton, Southampton SO17 1BJ, U.K.; Forensic Medicine and Science, University of Glasgow, Glasgow, UK; Health Research Institute, University of Limerick, Limerick, Ireland; Institute of Immunology and Immunotherapy, University of Birmingham, Birmingham, B15 2TT, U.K.; Centre of Membrane Proteins and Receptors (COMPARE), Universities of Birmingham and Nottingham, The Midlands, U.K.

**Author notes:** These authors have contributed equally.

## Abstract

**Objective:** Thrombotic complications and vasculopathy have been extensively associated with severe COVID-19 infection, however the mechanisms by which endotheliitis is induced remain poorly understood. Here we investigate vascular permeability in the context of SARS-CoV-2-mediated endotheliitis in patient samples and a vascular organoid model.

**Methods and Results:** We report the presence of the Spike glycoprotein in pericytes associated with pericyte activation and increased endothelial permeability in post-mortem COVID-19 lung autopsies. A pronounced decrease in the expression of the adhesion molecule VE-cadherin is observed in patients with thrombotic complications. Interestingly, fibrin-rich thrombi did not contain platelets, did not colocalize with tissue factor and have heterogenous levels of Von Willebrand factor, suggesting a biomarker-guided therapy might be required to target thrombosis in severe patients. Using a 3D vascular organoid model, we observe that ACE2 is primarily expressed in pericytes adjacent to vascular networks, consistent with patient data, indicating a preferential uptake of the S glycoprotein by these cells. Exposure of vascular organoids to SARS-CoV-2 or its antigens, recombinant trimeric Spike glycoprotein and Nucleocapsid protein, reduced endothelial cell and pericyte viability as well as CD144 expression with no additive effect upon endothelial activation via IL-1β.

**Conclusions:** Our data suggest that pericyte uptake of SARS-CoV-2 or Spike glycoprotein contributes to vasculopathy by altering endothelial permeability increasing the risk of thrombotic complications.

## Brief report

Severe acute respiratory syndrome coronavirus 2 (SARS-CoV-2)-associated vasculopathy and endotheliitis are common in severe cases of COVID-19^1^. Binding of the viral Spike glycoprotein to the host protein angiotensin-converting enzyme 2 (ACE2) is required for cellular entry, with different proteolytic reactions and co-receptors required for efficient virion infection^2,3^. Mechanisms of endotheliitis are poorly understood with conflicting evidence available as to whether SARS-CoV-2 infection damages 2D endothelial monolayers *in vitro*. Indeed, ACE2 expression in human endothelium is not sufficient for endothelial activation^4,5^ and infection of primary lung endothelial cells with SARS-CoV-2 results in limited infection^6,7^. Nevertheless, ACE-2-positive endothelial cells and disruption of intercellular junctions are observed in COVID-19 *post-mortem* lung sections^8^. Single cell RNAseq studies of healthy and fibrotic lungs have shown ACE2 expression on type II alveolar cells in healthy lungs and significant RNA expression on arterial vascular cells in fibrotic lungs^9^, a clinical complication observed in cohorts known to be vulnerable to COVID-19 infection^10^. In this letter, we show that SARS-CoV-2 antigens, namely the Spike glycoprotein and Nucleocapsid protein, induce pericyte activation and increase endothelial permeability, and this is associated with increased thrombotic complications in severe COVID-19 patients.

In the absence of direct evidence of endothelial viral uptake and subsequent damage, we reason that altered crosstalk between mural and endothelial cells (EC) promotes endotheliitis and regulates thrombosis. Pericytes are known to highly express ACE2 and regulate vascular integrity, permeability and blood flow^2,6,11^. We first assessed the presence of the Spike glycoprotein in the pericytes of lung tissues obtained *post mortem* from eight patients who died from COVID-19. All patients had comorbidities (Supplementary information). Spike glycoprotein was detected in the pericytes (NG2^+^ cells) of 4 patients, which also expressed high levels of the activation marker ICAM-1 (Figure 1A). As pericyte activation regulates endothelial permeability, we assessed the expression of VE-cadherin (CD144), a key regulator of vascular permeability, in non-ventilated COVID-19 patients. Compared to control lung sections from age-matched non-COVID-19 patients, CD144 expression was decreased in SARS-CoV-2 infected lungs, most markedly in patients with confirmed thrombosis (Figure 1B, C). These results suggest pericyte activation and compromised endothelial barrier function underlie the pathophysiology of severe SARS-CoV-2 infection.

**Figure 1:**
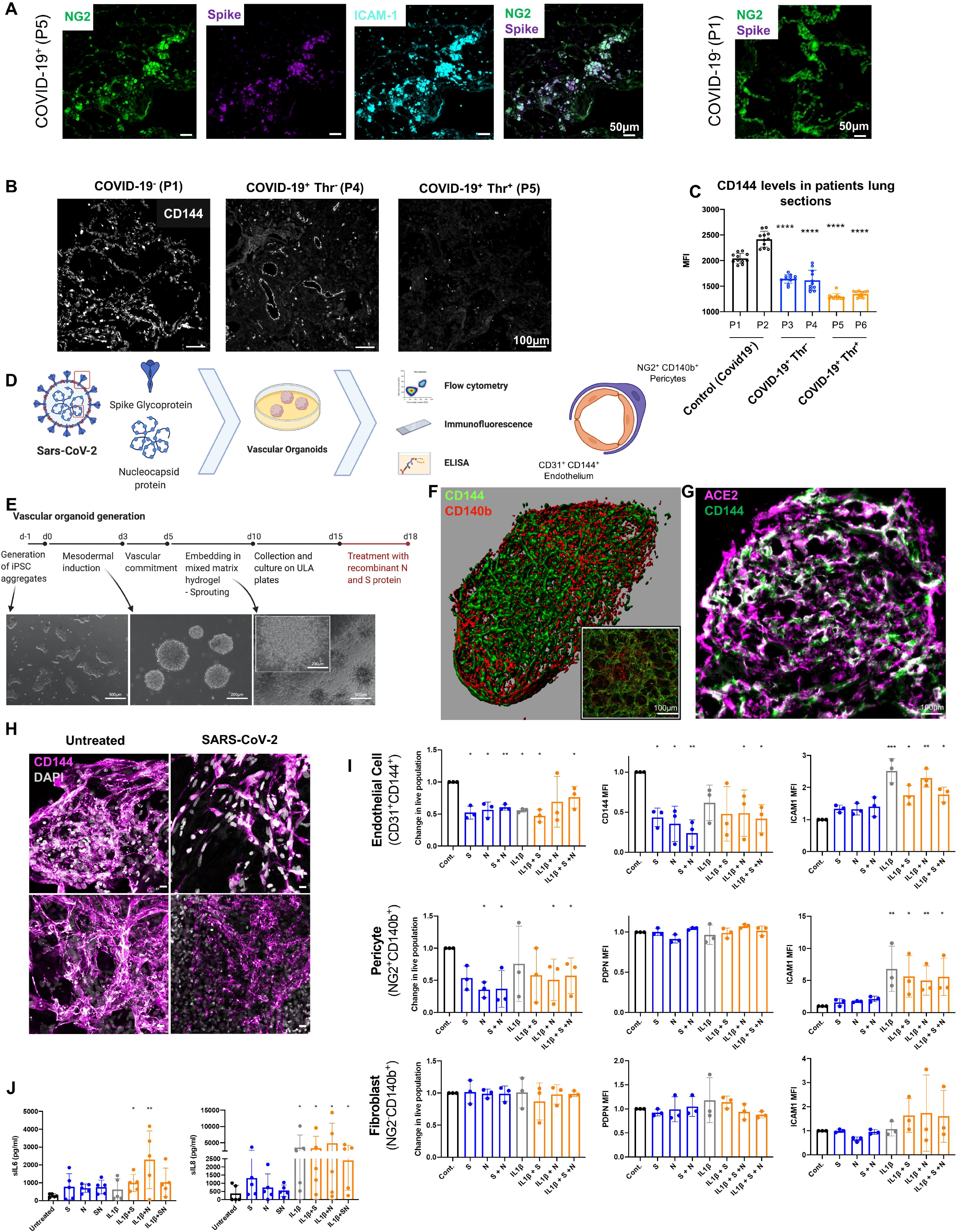
Increase in endothelial permeability and pericyte activation in COVID-19 lung sections and SARS-CoV-2 antigens-treated vascular organoids. (A) Immunofluorescence imaging of NG2^+^, Spike glycoprotein and ICAM-1 in formalin fixed and paraffin embedded lung from patient who died from COVID-19 and control COVID-19^-^ lung section. (B, C) Representative immunofluorescence imaging of CD144 (VE-cadherin) in lung sections (2 patients who died from non-respiratory-associated diseases (P1, P2), 2 COVID-19 patients without detectable thrombi (P3, P4 Thr^-^) and 2 patients with detectable microthrombi (P5, P6 Thr^+^)) as assessed by post-mortem histology. (C) Quantification of CD144 levels using ImageJ shows a significant reduction in CD144 expression in COVID-19^+^ patients, which is more notable in those with thrombosis. (D) SARS-CoV-2 England 2 virus (Wuhan) (M.O.I = 0.5) were added to the organoids for 48h. SARS-CoV2 trimeric Spike (S) and nucleocapsid (N) proteins (100nM) were added to vascular organoids model for 72h. (E) Vascular organoids were generated as reported by Wimmer *et al*. using a step wise differentiation of human induced pluripotent stem cells. First cells are committed to a mesodermal lineage as non-adherent aggregates, before embedding in a mixed matrix hydrogel which induces vascular sprouting. (F) On maturation, these samples are large 3D networks of branched endothelium (CD31^+^ CD144^+^) supported by pericytes (NG2^+^ CD140b^+^) and fibroblasts (NG2^-^ CD140b^+^). (G) Immunofluorescence imaging of ACE-2 and CD144^+^ positive cells in organoids (12µm organoid sections). (H) Immunofluorescence imaging of CD144 and nuclei (DAPI) in whole vascular organoids treated with SARS-CoV-2 (M.O.I= 0.5) for 48h or spike glycoprotein (S)(100nM) for 72h. (I) Activation of endothelial, pericytes and fibroblasts in the vascular organoids treated with Spike glycoprotein (S) and nucleocapsid (N), or a combination of both proteins for 72h in the presence or absence of IL1-β (20ng/ml) compared to control was measured by flow cytometry (Dako Cyan cytometer) after collagenase-digestion of vascular organoids. Median of fluorescence (MFI) of CD144 and ICAM-1 on CD31^+^ cells, ICAM-1 and podoplanin (PDPN) on CD140^+^NG2^+^ cells and CD140^+^NG2^-^ cells were measured by flow cytometry (n=3). (J) Detection of the levels of soluble IL-6 and IL-8 in the supernatant of organoids treated for 72h with viral antigens in the presence or absence of IL-1β by ELISA (n=4-5). One-Way ANOVA with multiple comparisons performed for each statistical test with significance at (* p = < 0.05, ** p = < 0.001).

In order to assess the direct role of SARS-CoV-2 on pericytes and endothelium, we generated 3D human vascular organoids ^12,13^ (Fig 1D, E) comprised of vascular endothelium (CD31^+^ CD144^+^), pericytes (CD140b^+^ NG2^+^), and fibroblast (CD140b^+^ NG2^-^) (Figure 1D,E, F) (Supplementary information). This model has been recently used to show that ACE2 antibody blockade limits viral replication *in vitro*^14^. ACE2 was primarily observed in pericytes adjacent to CD144^+^ cells, consistent with recent evidence of poor expression of ACE2 on endothelium^4^ (Figure 1G). Organoids were then exposed to i) live SARS-CoV-2 virus; ii) recombinant active trimeric spike glycoprotein (S) ^15,16^; iii) nucleocapsid protein (N) ^17^ or iv) a combination of both (S + N). Endothelial survival, permeability and activation, as well as pericyte and fibroblast survival and activation were assessed by flow cytometry and immunofluorescence (Figure 1D,E).

Treatment of vascular organoids with SARS-CoV-2 decreases CD144 expression on EC as observed by immunofluorescence, and trimeric S mimics this effect (Figure 1H). We next assessed the effect of S, N or S+N on endothelial, pericyte and fibroblast activation by flow cytometry. The decrease of CD144 was also observed following treatment with N and S+N Figure 1H, I) without significant changes in ICAM-1 (Figure 1I), CD62P or VCAM-1 expression on EC (data not shown for both), indicating that viral antigens do not directly activate the endothelium. Similarly, no activation of pericytes as measured by an increase in ICAM-1 expression was observed (Figure 1I). To assess whether endothelial activation alter EC response to S, N or S+N, we investigated the effect of SARS-CoV-2 antigens on cell activation in the presence of IL-1β, a chemokine known to activate endothelium and a critical component of the COVID-19 cytokine storm^18^. The SARS-CoV-2 antigen-dependent decrease in CD144 was not altered by the addition of IL-1β. Endothelial and pericyte cell activation by IL1-β, as assessed by the upregulation of ICAM-1, was not altered by the addition of SARS-CoV-2 proteins (Figure 1I). Neither recombinant SARS-CoV-2 antigens or IL-1β induced activation of fibroblasts in the vascular organoids, as measured by podoplanin expression. These results indicate that the SARS-CoV-2 antigens alter pericyte and endothelial survival and decrease CD144 on endothelial cells, increasing endothelial permeability.

Activated EC and pericytes release inflammatory cytokines such as IL-6 and IL-8, and higher levels are observed in severe COVID-19 patients ^19^. The levels of soluble IL-8 and IL-6 in the supernatant of treated organoids were modestly increased with SARS-CoV-2 antigens (Figure 1I). IL-1β increased IL-8 levels without further change by addition of SARS-CoV-2 antigens, showing that S and N have no additional effect on EC activation.

Pericyte activation and dysfunction increase endothelial cell procoagulant activity and endothelial permeability, as constitutive hypoplasia of pericytes increases VWF release, tissue factor expression and platelet adhesion^6^. Moreover, CD144 is crucial for endothelial stability and blockade of CD144 contributes to coagulopathy, particularly in the lung microvasculature^6,20,21^. We therefore assessed whether there is a link between CD144 expression and thrombosis in the lungs of fatal SARS-CoV-2 infected patients (n=8) (Figure 2A, B). Diffuse alveolar damage with extensive hyaline membrane rich in fibrin was observed in all patients (Figure 2A,B). Thrombi in the microvasculature was observed in 4 of eight lung autopsies of COVID-19 patients (Figure 2A). CD144 expression was decreased in all sections, and was almost undetectable in patients with diagnosed thrombosis (Figure 1B, 2B, C). Tissue factor was upregulated in all lung sections, independently of thrombosis (n=9), suggesting no direct correlation between thrombosis and upregulation of tissue factor. Thrombosis was associated with increased platelet recruitment and accumulation in 2 patients as single platelets or in microaggregates, without evidence of fibrin deposition on these thrombi in the alveolar capillaries while fibrin rich thrombi did not contain platelets, which were localized on the endothelium (Figure 2B). It is possibly that the presence of procoagulant phosphatidylserine-positive platelets observed in severe COVID-19 patients support the generation of fibrin-rich thrombi ^22^. VWF and platelets were observed in some but not all patients diagnosed with thrombosis, suggesting distinct thrombus composition (P6). Moreover, megakaryocytes were also observed in 3 patients, with enrichment around hyaline membrane^23,24^. We further compared the expression of CD144 in Middle East respiratory syndrome coronavirus (MERS-CoV)- and rhinovirus-to SARS-CoV-2-infected lung specimen. A decrease in CD144, increased fibrin deposition and platelet recruitment in the microvasculature is observed in a MERS-infected lung associated with thrombosis (Figure 2D) whereas CD144 expression was more evident in rhinovirus-infected lung negative for thrombosis, fibrin deposition and platelet recruitment (Figure 2E). These results suggest that a decrease in CD144 is associated with higher risk of thrombosis in the lung microvasculature during SARS-CoV-2 and MERS-CoV infections. Further studies with larger cohorts of viral infections are necessary to confirm these observations.

**Figure 2:**
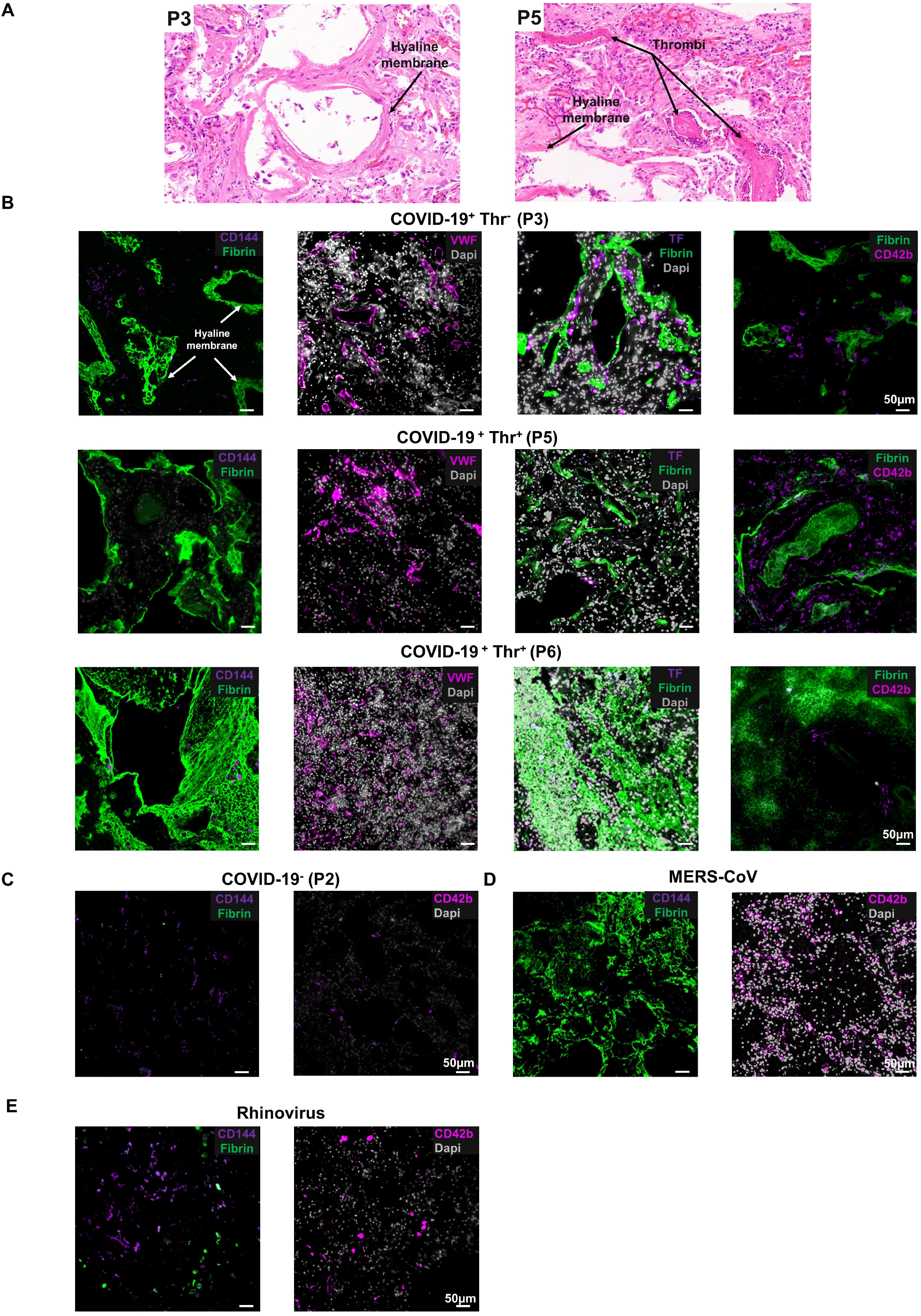
Endothelial permeability and thrombosis in COVID-19 lungs. (A) H&E staining of lung sections from patients who died from COVID-19 with or without evidence of thrombosis. (B) Immunofluorescence imaging of CD144, Von Willebrand Factor (VWF), tissue factor (TF), platelets (CD42b) and fibrin in formalin fixed and paraffin embedded lung sections from control, COVID-19, Rhinovirus and MERS-CoV patients. Nuclei/DNA were stained with Dapi. Images was captured using Epi fluorescence microscope and slide scanner Axio Scan. Z1.

In summary, using a vascular organoid model, we show that SARS-CoV-2 antigens directly alter endothelial permeability by decreasing CD144 expression and have a detrimental effect on endothelial and pericyte populations. The presence of comorbidities might account for increased endothelial permeability and favour the passage of the virus across the endothelial barrier and pericyte infection altering endothelial permeability, exacerbating endotheliitis, and ultimately contributing to the thrombotic complications which are fatal in severe infections. This effect appears independent of classical endothelial activation, however the elevated inflammatory cytokines such as IL-1 β could contribute to the endotheliitis. Dual therapies regulating endothelial activation and permeability is likely to be beneficial in severe patients to limit endotheliitis and thrombosis. However, as thrombus composition is not homogenous among patients, biomarker-guided strategies might be required to target thrombosis in COVID-19 patients.

## Materials and Methods

### Patients

*Post mortem* formalin-fixed and paraffin-embedded lung sections from eight COVID-19, one MERS-CoV, one rhinovirus and 2 control (non-respiratory-associated diseases) patients. Ethics for patient tissue sections were approved by the Health Research Authority (HRA) with an NHS (National Health Service) REC (Research Ethics Committee) approval issued by North East-Newcastle and North Tyneside 1 (19/NE/0336). Ethical approval was granted. COVID-19 samples were obtained from COVID-19 cases (pre-hospital and hospital deceased patients), with time from symptoms to death ranging from 0-36 days, and ages ranging from 59-89 years old. Patients were not on mechanical ventilation and all had comorbidities. Patients comorbidities are shown in the table below.

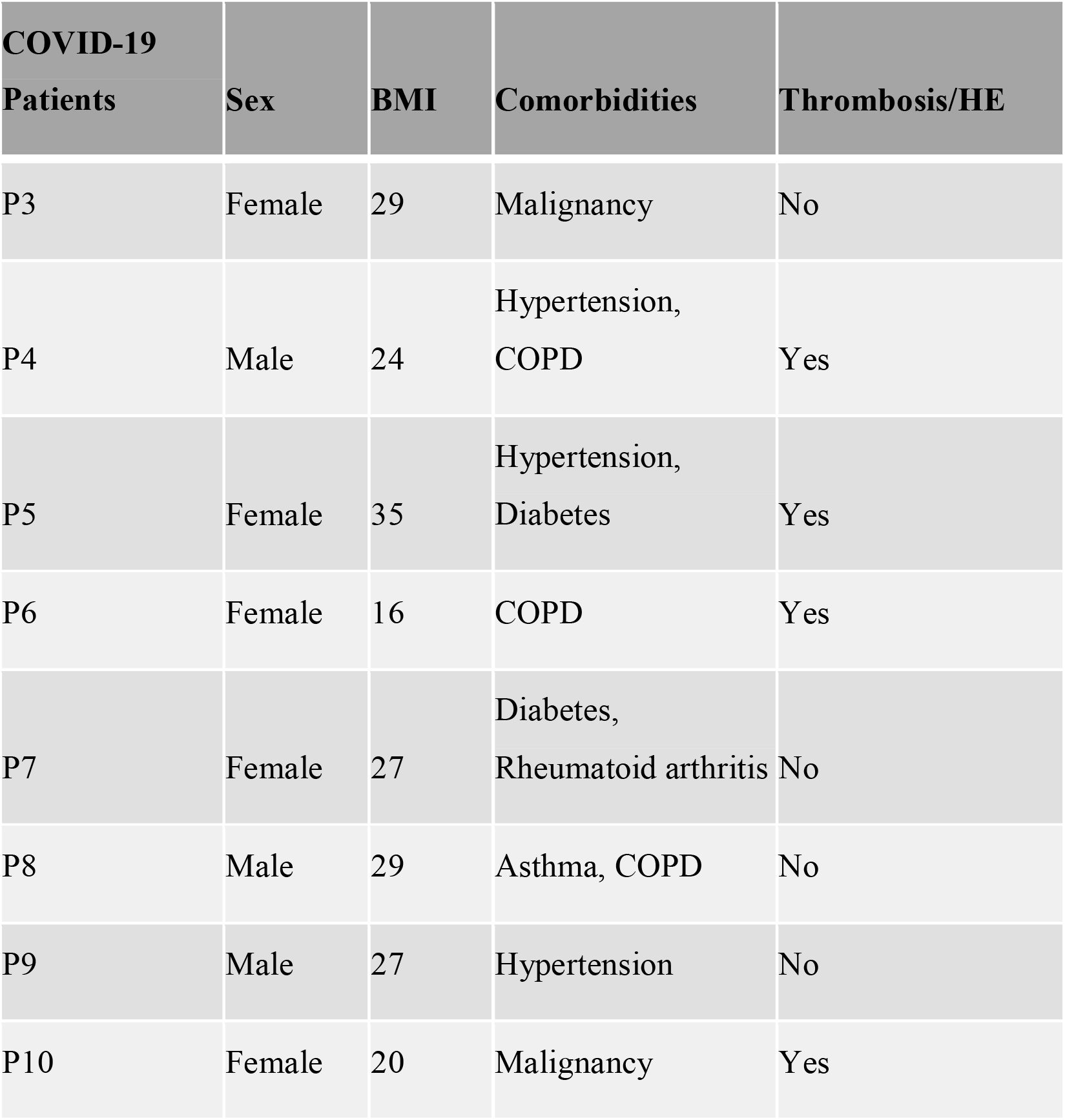

### iPSC culture and organoid generation

Human induced pluripotent stem cells were obtained from Gibco (Thermo) and cultured on GelTrex (Thermo) coated 6-well plates in StemFlex medium (Thermo). Cells were passaged using an EDTA dissociation method, and routinely karyotyped as described by Khan *et al*. Vascular organoid generation was performed in a manner similar to that described by Wimmer *et al*. Briefly, cells were dissociated and replated on Ultra Low Attachment 6-well plates (Corning) in StemFlex supplemented with RevitaCell (Thermo) overnight before commencement of differentiation protocol. On day 0 (d0), cells were collected from Ultra Low Attachment plates and spun down at 500G before resuspension in Phase I media, which was comprised of APEL2 (Stem Cell Technologies) media supplemented with CHIR90921 (12uM), and BMP4 (Thermo), FGF2, and VEGFA at 50ng/mL (Stem Cell Technologies). Cells were incubated at 37 degrees in 5% CO_2_ for 3 days, before pelleting by gravitation and resuspension in Phase II medium. Phase II medium was composed of APEL2 medium, VEGFA at 100ng/mL, and FGF2 at 50ng/mL. On day 5 of the protocol, mesodermal blasts were embedded in a mixed matrix hydrogel composed of 60% Collagen Type I (VitroCol – Advanced Biomatrix) and reduced Growth Factor Matrigel (Corning), and incubated in phase II medium supplemented with 15% FBS. Fresh media was added on day 8, and sprouted cultures were isolated from the hydrogel at day 10 by scraping and centrifugation. Collected organoids were then cultured individually in 96 well Ultra Low attachment plates (Corning) before treatment at day 15. For imaging, fixation was performed in 10% formalin. For flow cytometry organoids were dissociated in Collagenase Type II (200 U/mL).

### Treatment vascular organoids with SARS-CoV-2 virus or antigens

SARS-CoV-2 England 2 virus (Wuhan) was a kind gift from Christine Bruce, PHE. Recombinant trimeric Spike glycoprotein (S) was produced as previously described ^15,16^. Nucleocapsid (N) protein was produced and purified as described ^17^. Vascular organoids were treated for 48h with SARS-CoV-2 virus (M.O.I = 0.5). S and N (100nM) were added to vascular organoids for 72h in the presence and absence of IL-1β (20ng/ml) (Peprotech). Soluble IL6 (sIL6) and sIL8 were measured in the supernatant by ELISA (Peprotech).

### Organoids and Lung sections staining

#### Whole organoid staining

Fixed, whole organoids were prepared for immunofluorescence by first incubating fixed samples overnight in blocking buffer (2% goat serum, 1% bovine serum albumin) supplemented with Triton X100, Tween, and Sodium Deoxycholate. Samples were then incubated in blocking buffer and primary antibody against CD144 overnight, before sequential 5 minutes washes in PBS-Tween and a 2 hours incubation in PBS-Tween with secondary antibody (AlexaFluor 647 and DAPI). Upon labelling samples were washed again in phosphate buffer saline (PBS)-Tween, before mounting in 0.5% low melting point Agarose (Fisher Scientific) in an Ibidi 8-well slide (Ibidi). Samples were then subject to serial dehydration in ethanol (30%, 50%, 70%, 100%) before clearing in ethyl cinnamate and imaging using Airyscan confocal microscopy.

#### Organoid sections staining

Vascular organoids were frozen in optimum cutting temperature (OCT) compound (Tissue-Tek, The Netherlands),12µm sections were performed and blocked with PBS containing 5% BSA and 10% goat serum. Primary antibodies against ACE-2 (Thermofischer) and CD144 (Thermofischer) were used for immunofluorescence.

#### Lung sections staining

For paraffin sections, following rehydration and antigen retrieval, lung sections were treated with H2O2 3% for 15 minutes and blocked with PBS containing 5% bovine serum albumin and 10% goat serum for 1h. Antibodies against Neural/glial antigen 2 (NG2) (ebiosciences), Spike glycoprotein (Sinopharma), Intercellular Adhesion Molecule 1 (ICAM-1, CD54) (Biolegend), CD144 (VE-cadherin) (Thermofischer), ACE-2 (Thermofisher), VWF (Agilent), Tissue Factor TF (Abcam), platelet CD42b (Abcam) and fibrin (Merck Life Science) were incubated overnight at 4°C. Secondary antibodies were added for 1h at room temperature. Nuclei was staining using Dapi. Lung autofluorescence was quenched using commercial kit (Vector laboratories) and slides mounted using ProLong Gold Antifade Moutant (Life Technologies). Sections were images using Epi fluorescent microscopy or Zeiss Axio Scan.Z1 microscope and analysed using ZEN software and image J.

### Flow cytometry

Collagenase-dissociated organoids were blocked with PBS-FBS 10% for 20 min on ice. Cells were stained with anti-CD144-PEcy7, anti VCAM-PEcy5, anti CD62P-Pecy.5, anti podoplanin-FITC, anti CD140-PE, anti CD31-APCcy7 (all from Biolegend), anti ICAM-1-Biotin followed by Streptavidin PE-CF594 (BD), anti NG2-APC (Bio-Techne Ltd), Live/Dead Fixable Aqua Dead Cell Stain (Thermofischer) for 20 minutes on ice. Cells were fixed and acquired by flow cytometry (Beckman Coulter).

### Data analysis

All data were presented as means □±□s.d. The significant difference groups were analyzed using either a One-Way ANOVA with multiple comparisons or a Kruskal-Wallis Test with multiple comparisons as indicated in figure legends using Prism 7 (GraphPad Software Inc, USA).

## Data Availability

Please contact corresponding authors re any data requests.

## Acknowledgements

AOK is a Henry Wellcome fellow (218649/Z/19/Z). JR is a British Heart Foundation Intermediate Fellow (FS/IBSRF/20/25039). This research was supported by BHF Accelerator Awards to JR and AOK (AA/18/2/34218). This research was funded, in whole or in part, by the Wellcome Trust (218649/Z/19/Z), British Heart Foundation (AA/18/2/34218) and COMPARE. A CC BY or equivalent licence is applied to AAM arising from this submission, in accordance with the grant’s open access conditions. M.C. was supported by the National Institute for Allergy and Infectious Diseases through the Scripps Consortium for HIV Vaccine Development (CHAVD) (AI144462), the University of Southampton Coronavirus Response Fund. M.C. was also supported by the International AIDS Vaccine Initiative (IAVI) through grant INV-008352/ OPP1153692 and the IAVI Neutralizing Antibody Center through the Collaboration for AIDS Vaccine Discovery grant INV-008813/OPP1196345, both funded by the Bill and Melinda Gates Foundation.

## Authorship

Contribution: AOK and JR designed research, performed research, analyzed data and wrote the paper; JSR, JHB and MC performed research; MLN, JDA and MC contributed vital new reagents; EY, PGM, GT, AGR contributes to clinical information and samples collection; ZS and MP contributed vital new reagents, performed experiments and analysed data; AF contributed vital new reagents and contributed to data analysis. All authors read and approved the paper.

## Conflict-of-interest disclosure

The authors declare no competing financial interests.

## Notes

### Competing Interest Statement

The authors have declared no competing interest.

### Author Declarations

Ethics for patient tissue sections were approved by the Health Research Authority (HRA) with an NHS (National Health Service) REC (Research Ethics Committee) approval issued by North East- Newcastle and North Tyneside 1 (19/NE/0336). Ethical approval was granted.

